# Dialectical Behavior Therapy Skills Training as a Brief Intervention for Cigarette Smoking by Patients with Cancer: A Scoping Review and Narrative Synthesis of Related Literature

**DOI:** 10.1101/2024.10.13.24315419

**Authors:** Marcia H. McCall, Charlotte T. Boyd, Nicole D. Kerr, Stephanie S. Daniel, Erin L. Sutfin

## Abstract

**Objective:** Novel behavioral interventions are needed for patients with cancer who smoke cigarettes. Standard tobacco treatment may not effectively address the psychological distress and/or emotion dysregulation that makes quitting smoking difficult for many patients. Dialectical Behavior Therapy – Skills Training (DBT-ST) has demonstrated efficacy as a brief intervention for managing emotions and stress across varied populations, but has not been adapted for patients with cancer who smoke. To determine its suitability for this population, we conducted a scoping review of brief DBT-ST with similar populations: people with substance use, breast cancer, or emotion dysregulation.

**Methods:** We followed PRISMA-ScR (Preferred Reporting Items for Systematic reviews and Meta-Analyses extension for Scoping Reviews) guidelines. Studies were restricted to English-language publications of DBT-ST as a brief intervention of 20 or fewer sessions. We found 26 publications representing 23 research studies, extracted study details, and narratively synthesized the results.

**Results:** The 23 studies included 12 quasi-experimental designs, seven pilot randomized controlled trials (RCTs), and four RCTs. All studies found at least one improvement in a main outcome following DBT-ST intervention, with results maintained at follow-up. Qualitative outcomes indicated high satisfaction with DBT-ST and good retention. Studies recruited diverse participants, with some far exceeding population averages. Over half of studies included only females or males. We found considerable heterogeneity across studies in intervention design, testing, and measurement.

**Conclusion:** DBT-ST as a brief intervention for people with substance use, cancer, or emotion dysregulation demonstrates sufficient positive outcomes to adapt this approach for patients with cancer who smoke cigarettes.

## Background

### Rationale

Patients with cancer who smoke cigarettes experience more treatment failures and complications and are less likely to survive cancer than patients who stop smoking or have never smoked.^1,2^ In contrast, the benefits of not smoking during or after cancer treatment are well-established, among them improved survival, better surgical outcomes, lower likelihood of recurrence or new tumor sites, less fatigue, better cognitive processing, and increased psychological well-being compared to continuing to smoke.^3,4^ Although a cancer diagnosis can be a catalyst for addressing smoking, 12.7% of patients with cancer continue to smoke cigarettes.^5,6^

The standard of care tobacco counseling recommended for patients with cancer employs motivational and behavioral strategies to help patients manage withdrawal, plan for situations that may trigger the desire to smoke, connect their motivation to quit with their values and goals, and build confidence to quit.^7^ However, tobacco counseling is often ineffective with these patients, for whom smoking can be a chronic, relapsing addiction.^8,9^ The unique psychological mechanisms of patients with cancer may explain the failure of tobacco counseling to help them change smoking behaviors. First, for people who smoke cigarettes at the high levels causing cancer, smoking is a tenacious, entrenched habit usually functioning as a primary coping mechanism for managing life stress and negative emotions.^10,11^ Smoking provides relaxation and comfort (in part by relieving nicotine cravings) during disruptive life events such as cancer diagnosis and treatment. Second, patients with cancer who smoke are more likely to have experienced persistent childhood trauma, a strong predictor of both heavy smoking and cancer, with rates disproportionately higher among patients who are Black or Latine.^12–16^ Childhood traumatic experiences frequently contribute to distress intolerance and emotion dysregulation that may be worsened by the added burden of dealing with cancer.^13,15,16^ Emotion dysregulation is a trans-diagnostic destabilizing dynamic for people with psychological distress, including patients with cancer.^17–20^ Third, having cancer and undergoing treatment may induce or exacerbate stress, anxiety, depression, hopelessness, and fatalistic beliefs.^10,11,21^ These negative affect states predict a lower likelihood of quit attempts,^21^ whereas the ability to tolerate stress increases the likelihood of quitting smoking and remaining smoke free for at least one year.^22^ Behavioral interventions addressing the psychological concerns of patients with cancer who smoke are critically needed.^23^

Substantial evidence supports that standard of care tobacco counseling methods have been less effective for patients who continue to smoke following cancer diagnoses.^8,9^ A promising behavioral counseling alternative for this patient population is Dialectical Behavior Therapy (DBT), an evidence-based psychotherapeutic approach developed specifically for people needing help managing stress and negative emotions. *Dialectic* means the synthesis of opposites. In DBT, the dialectic is the synthesis of *change* in emotions and relationships and *acceptance* of distress and self.^24^ DBT emerged in the late 1970’s as an effective treatment for people with borderline personality disorder (BPD), characterized by emotional volatility, low stress tolerance, and suicidal ideation, following the failure of cognitive-behavioral therapy to improve this condition.^25^ As with smoking and cancer, pervasive childhood trauma strongly predicts BPD.^26^ DBT helps people with BPD and other conditions, such as eating disorders, gain proficiency in distress tolerance, emotion regulation, interpersonal effectiveness, and mindfulness.^25^ DBT can be tailored for greater acceptability by patients from varied cultural backgrounds.^25,27,28^

The full DBT program involves one to two years of weekly group meetings, individual therapy sessions, and telephone counseling, facilitated by trained behavioral health clinicians.^25^ This structure limits access for many people who might benefit from DBT, such as patients with cancer. Accordingly, during the past two decades, researchers developed and tested brief DBT interventions as short as four sessions using content and processes from DBT’s central component, Skills Training (DBT-ST). These DBT-ST adaptations provided effective brief treatments for populations similar to patients with cancer who smoke, including people with substance use disorders, patients with breast cancer experiencing psychosocial stress, and people with emotion dysregulation.^27,29–34^

### Objective

A review of brief DBT-ST interventions (20 or fewer sessions) has not been conducted but could yield valuable information regarding intervention design and efficacy across populations. For emerging and evolving areas, a scoping review is appropriate to map the literature and to identify gaps for future direction.^35^ Accordingly, we conducted a scoping review to locate, examine, and narratively synthesize studies of DBT-ST adaptations as brief interventions for adult populations with substance use disorders, breast cancer and psychosocial stress, and emotion dysregulation, in consideration of potentially adapting DBT-ST for patients with cancer who smoke cigarettes.

## Methods

We conducted a scoping literature review following PRISMA-ScR (Preferred Reporting Items for Systematic reviews and Meta-Analyses extension for Scoping Reviews) guidelines to locate studies of DBT-ST adapted as brief interventions for the selected populations.^36^

### Eligibility Criteria

We included peer-reviewed journal articles of DBT-ST as a standalone brief intervention, delivered to individuals and/or to groups of adults (aged 18 and older) in any outpatient or community setting. Our conditions of interest – to align with our target population of patients with cancer who smoke cigarettes – were substance use disorders, cancer diagnoses, and emotion dysregulation. We excluded studies of DBT-ST for people diagnosed with personality, behavioral, or attention disorders; with serious mental illnesses such as bipolar disorder; or with suicidal or non-suicidal self-injury behaviors. We included any quantitative or mixed-methods research design that reported patient- or group-level outcomes. Interventions needed to be sufficiently described to determine whether formal DBT skills training had occurred. To align with the number of sessions potentially feasible for patients with cancer, we included DBT-ST interventions of no more than 20 sessions. Studies with concurrent wraparound DBT components – telephone calls, individual therapy – were excluded, but studies with non-DBT components (usually specific to the population, e.g., case management for justice-involved veterans) were included. We required full text availability and publication in the English language. Studies were not restricted by date or country of origin.

### Information Sources, Searches, and Selection of Studies

In March and April 2024, we searched the CINAHL, ProQuest Psychology, PsycINFO, and PubMed databases, as well as Google Scholar. We searched all fields using the keywords Dialectical Behavior Therapy, cancer, emotion dysregulation, and substance use and their variants. For example, our PubMed search syntax was (“dialectical behavior therapy”[All Fields]) AND ((“cancer”[All Fields]) OR (“emotion dysregulation”[All Fields]) OR (“substance use”[All Fields])). Eligibility criteria and search results were uploaded to Covidence, a web-based collaboration software platform that streamlines the production of systematic and other literature reviews.^37^ Three reviewers (MM, CB, NK) examined titles and abstracts generated from each database to locate studies meeting eligibility criteria, with agreement by two reviewers sufficient to advance a publication to full-text review. We conducted full-text review, requiring agreement by all three reviewers for study inclusion. Differences were resolved during four consecutive weekly meetings of the three reviewers. To locate studies missed by the search process, we examined reference lists of studies meeting inclusion criteria and of DBT literature reviews found during the searches, again requiring agreement by all three reviewers for study inclusion.

### Data Charting, Items, and Synthesis

We performed data extraction and charting directly from publications to Microsoft Excel tables. We extracted research design; study location; population including demographic characteristics and condition(s) under study; specifics about the intervention and comparison conditions such as number of sessions and DBT-ST topics addressed; measures; and individual- and/or group-level outcomes. As most studies were quasi-experimental or pilot studies, we did not conduct critical appraisal of studies. We performed a narrative synthesis of the extracted results.

## Results

After screening the titles and abstracts of 2738 publications and performing full text review of 50 studies, we located 26 publications that met our inclusion criteria, representing 23 unique studies with four publications from a single pilot RCT ^30,38–40^ (see *Figure 1*. *PRISMA 2020 flow diagram*). Data results are presented in *Table 1. Research Design, Population, Intervention, and Outcomes*, followed by our narrative synthesis of the data.

**Figure 1.**
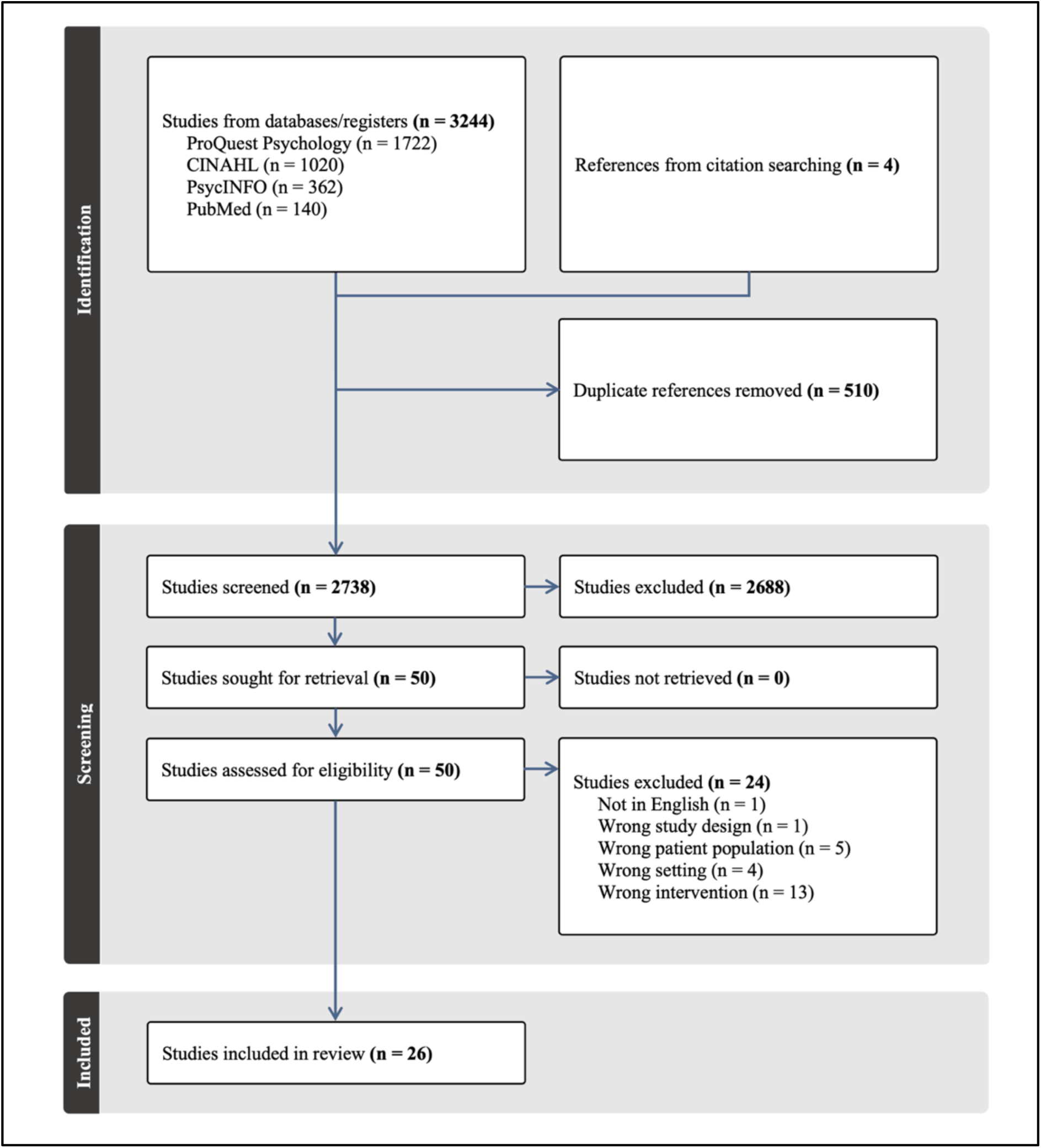
PRISMA 2020 Flow Diagram

**Table 1.** Research Design, Population, Intervention, and Outcomes.

| Article, Research Design, Country | Population, Condition, Demographics | Intervention, Comparison | Primary Outcomes |
| --- | --- | --- | --- |
| **Cancer** |  |  |  |
| Anderson et al. (2013)<br>Quasi-experimental<br>United States (Wisconsin) | N = 14<br>100% completion rate (implied)<br>Breast cancer<br>100% female<br>Race/ethnicity not reported<br>Age range: 37 - 64 | DBT-ST individual and group sessions, pre-post, no comparison<br>DBT-ST modules: all<br>DBT-ST sessions: 8 (session length not reported) | Pre vs. post: improvement in distress level ($p < .001$)<br>Participants reported qualitative improvement in insight and behavior including ability to cope, recognize stress triggers and use skills |
| Faraj (2015)<br>Pilot RCT<br>Iran (Tehran) | N = 30<br># Non-completers not reported<br>Breast cancer<br>100% female<br>Race/ethnicity and age not reported | DBT-ST group vs. treatment-as-usual<br>DBT-ST modules: all<br>DBT-ST sessions: 10 x 90 minutes each | DBT-ST vs. treatment as usual: improvements in distress level ($p < .001$); coefficient of determination - 48.9% of reduction in distress is explained by intervention<br>DBT-ST vs. treatment as usual: improvements in life expectancy given cancer ($p = .023$); coefficient of determination - 17.6% of increase in life expectancy is explained by intervention |
| **Emotion Dysregulation** |  |  |  |
| Afshari & Hazani (2020)<br>RCT<br>Iran (Isfahan province) | N = 68<br>Completed case analysis<br>N = 63 (32 DBT, 31 CBT)<br>Generalized anxiety disorder<br>59% female<br>100% White<br>Age range: 18 - 45 | DBT-ST group vs. CBT group<br>DBT-ST modules: all<br>DBT-ST and CBT sessions: 16 x 60 minutes each | DBT-ST *and* CBT: improvements in emotion regulation, mindfulness, depression and anxiety ($p < .05$), maintained at 3-month follow-up, with greater improvements for emotion regulation and mindfulness in the DBT-ST group and for depression and anxiety in the CBT group |
| Babaheydari et al. (2024)<br>RCT<br>Iran (Fars) | N = 40<br>100% completion rate (implied)<br>Generalized anxiety disorder<br>0% female<br>Race/ethnicity not reported<br>Age range: 20-30 | DBT-ST group vs. waitlist control<br>DBT-ST modules: all<br>DBT-ST sessions: 10 x 90 minutes each | DBT-ST vs. control: improvements in emotion regulation and anxiety ($p < .0001$) maintained at 3-month follow-up. Improvements consistent across all subscales of emotion regulation instrument. |
| Davarani & Heydarinasab (2019)<br>Quasi-experimental<br>Iran (Tehran) | N = 20<br>100% completion rate (implied)<br>Emotion dysregulation<br>100% female<br>Race/ethnicity not reported<br>M age: 21.5 (SD 2.5)<br>Age range: 18-25 | DBT-ST group, pre-post vs. treatment-as-usual<br>DBT-ST modules: all<br>DBT-ST sessions: 8 x 120 minutes | DBT-ST vs. control: improvements in six of seven emotion regulation subscales ($p < .01$) except for "carrying out purposeful behavior" |
| Harley et al. (2008)<br>Pilot RCT<br>United States (Massachusetts) | N = 24<br>Completed case analysis<br>N=19 (10 DBT-ST, 9 waitlist)<br>Treatment-resistant depression<br>75% female<br>83% White, 17% other<br>M age=41.8 | DBT-ST group vs. waitlist control<br>DBT-ST modules: all<br>DBT-ST sessions: 16 x 90 minutes each | DBT-ST vs. waitlist: improvement in depressive symptoms - clinician-rated symptoms ($p < .05$, $d = 1.45$) and self-report symptoms ($p < .01$, $d = 1.31$), both maintained at 6-month follow-up (but only 8 participants included in follow-up) |
| Khodabakhshi-Koolae et al. (2024)<br>Quasi-experimental<br>Iran (Tehran) | N = 30<br>100% completion rate<br>Emotion dysregulation and death anxiety<br>100% female<br>Race/ethnicity not reported<br>M age=63.01 (SD 3.16), age range 60-75 | DBT-ST group vs. treatment-as-usual<br>DBT-ST modules: all<br>DBT-ST sessions: 10 x 70-75 minutes each | DBT-ST vs. control: improvements in emotion regulation ($p=.001$, $\eta^2=.357$) and death anxiety ($p=.0001$, $\eta^2=.531$). Follow-up not possible due to COVID-19 restrictions |
| Lee & Arora (2022)<br>Quasi-experimental<br>United States (Maryland) | N = 59<br>92% completion rate (attending at least 75% of sessions)<br>Mental health disorders - anxiety, depression, eating<br>75% female<br>23.7% Black, 5.1% Hispanic/Latine, 45.8% White, 15.3% Asian, 10.2% multiracial<br>M age=21.3 (SD 2.95), age range 18-31 | DBT-ST group vs. treatment-as-usual<br>DBT-ST modules: distress tolerance, emotion regulation, mindfulness<br>DBT-ST sessions: 4 x 90 minutes each | Pre vs. post: improvements in emotion regulation ($p<.001$, $d=-0.66$), mindfulness ($p<.001$, $d=0.54$), overall distress ($p=.005$, $d=-0.41$), and resilience ($p<.001$, $d=0.70$)<br>Outcomes continued to improve at 1-month and 3-month follow-up, but follow-up rates were modest (29% and 22%, respectively)<br>Qualitative evaluation of acceptability: 92% attended at least 3 of 4 sessions, 93% gained useful skills, 91% were better able to cope, and 70% saw symptoms decrease |
| Neacsiu et al. (2014)<br>Pilot RCT<br>United States (Washington) | N = 48<br>ITT analysis with N=44<br>Emotion dysregulation<br>66% female<br>7% Hispanic/Latine, 93% White<br>M age: 35.6 (SD 12.4), age range 19-70 | DBT-ST group vs. activities-based support group, with rolling admissions at weeks 1, 2, and 9<br>DBT-ST modules: all<br>DBT-ST and support group sessions: 16 x 2 hours | DBT-ST vs. support group: improvements in emotion regulation for both groups, but DBT-ST improved more rapidly ($d=1.86$), with skill use frequency ($d=1.02$) explaining 62.3% of change in emotion regulation<br>DBT-ST vs. support group: greater decrease in anxiety ($d=1.37$) with skills use explaining 47.6% of change, but no difference for depression ($d=0.73$)<br>Feasibility: difference in dropout rates between groups not significant. DBT-ST participants more likely to attribute improvements in depression and anxiety to DBT-ST vs. support group participants to group activities ($p<.01$) |
| Neacsiu et al. (2018)<br>Pilot RCT<br>Secondary analysis of Neacsiu et al. (2014)<br>United States (Washington) | N = 48<br>ITT analysis with N=44<br>Emotion dysregulation in anger, shame, disgust<br>66% female<br>7% Hispanic/Latine, 93% White<br>M age: 35.6 (SD 12.4), age range 19-70 | DBT-ST group vs. activities-based support group, with rolling admissions at weeks 1, 2, and 9<br>DBT-ST modules: all<br>DBT-ST and support group sessions: 16 x 2 hours | DBT-ST vs. support group: improvements in anger suppression ($p<.001$, $d=0.93$) and distress ($p<.001$, $d=1.04$), indirectly mediated by improvements in emotion regulation<br>DBT-ST and support group: both treatments reduced shame and disgust, and neither treatment affected anger expression |
| Southward, Eberle, & Neacsiu (2022)<br>Pilot RCT<br>Secondary analysis of Neacsiu et al. (2014)<br>United States (Washington) | *N* = 19<br>ITT analysis (original *N*=24)<br>Emotion dysregulation<br>68% female<br>5% Hispanic/Latine, 95% White<br>*M* age: 31.7 (*SD* 8.95) | DBT-ST group vs. activities-based support group, with rolling admissions at weeks 1, 2, and 9<br>DBT-ST modules: all<br>DBT-ST and support group sessions: 16 x 2 hours | Pre vs. post: improvements in skills use (*M* weekly increase=0.14 skills, 95% *CI* [.10, .18], *p*<0.01). More skills were used on days with greater stress and anxiety (*p*<0.01), which predicted next-day decreases in stress and anxiety (*p*<0.01). Higher effectiveness of skills use reported when experiencing intense negative affect (*p*<0.01)<br>Pre vs. post: no improvement in self-reported effective use of DBT-ST skills or in anxiety, depression, or stress |
| Wyatt et al. (2022)<br>Pilot RCT<br>Secondary analysis of Neacsiu et al. (2014)<br>United States (Washington) | *N* = 48<br>ITT analysis with *N*=44<br>Emotion dysregulation<br>66% female<br>7% Hispanic/Latine, 93% White<br>*M* age: 35.6 (*SD* 12.4), age range 19-70 | DBT-ST group vs. activities-based support group, with rolling admissions at weeks 1, 2, and 9<br>DBT-ST modules: all<br>DBT-ST and support group sessions: 16 x 2 hours | Within-person: at each time point, lower emotion dysregulation predicted by greater skills use (*B*=-0.52, *p*<0.001), mindfulness (*B*=-0.72, *p*<.001), and perceived control (*B*=-0.19, *p*<.001). Results did not hold across time points, nor did between-person variability mediate the effects |
| Rivzi & Steffel (2014)<br>Quasi-experimental<br>United States (New Jersey) | *N* = 24<br>ITT analysis with 3 non-completers<br>Emotion dysregulation<br>88% female<br>12% Black, 71% White, 17% Asian<br>Age range: 18 - 29 | DBT-ST emotion regulation + mindfulness group vs. DBT-ST emotion regulation-only group<br>DBT-ST modules: emotion regulation, mindfulness<br>DBT-ST sessions: 8 x 2 hours | DBT-ST emotion regulation + mindfulness group vs. DBT-ST emotion regulation group: no significant differences between groups<br>Pre-post combined groups: significant improvements in emotion regulation (*p*<.001), stress (*p*<.001), mindfulness (*p*<.001), positive affect (*p*<.001), negative affect (*p*<.001), and DBT skills use (*p*<.001) |
| Robins, Roberts, & Sarris (2018)<br>Quasi-experimental<br>Australia (South Australia, Victoria) | *N* = 74<br>Intervention (*N*=17) 100% completion, control (*N*=57) 65% completion<br>ITT analysis with 3 non-completers<br>Psychological distress and burnout<br>Across groups:<br>93%-94% female<br>12% Black, 71% White, 17% Asian<br>*M* age=30.84-31.53 (*SD* 6.37,6.60), age range 23-52 | DBT-ST group vs. treatment-as-usual<br>DBT-ST modules: all<br>DBT-ST sessions: 8 x 2 hours | DBT-ST vs. control: improvements in psychological distress (*p*=0.000, *d*=-1.68, 95% *CI* [-0.99, -2.32]), emotional stability (*p*=0.022, *d*=-0.71, 95% *CI* [-0.10, -1.30]), and mindfulness (*p*=0.001, *d*=1.39, 95% *CI* [0.72, 2.01]), with outcomes maintained at 6-month follow-up<br>Qualitative results found 94.5% of DBT-ST group found skills and group useful and would recommend to others, with themes supporting increased wellbeing, effectiveness of group structure, and usefulness of skills. |
| Shojaei-Fadafen, Jajarmi, & Mahdian (2022)<br>Quasi-experimental<br>Iran (North Khorasan) | N=30<br>Psychological distress and depression<br>100% female<br>Race/ethnicity not reported<br>Most frequent age range=30-39 (67%) | DBT-ST group vs. waitlist control<br>DBT-ST modules: distress tolerance, emotion regulation, mindfulness<br>DBT-ST sessions: 8 x 90 minutes each | DBT-ST vs. control: improvements in depression ($p=0.001$), irrational beliefs ($p=0.003$), and psychological wellbeing ($p=0.002$), with outcomes maintained at 3-month follow-up |
| Uliaszek et al. (2015)<br>Pilot RCT<br>Canada (Ontario) | $N = 54$<br>ITT analysis with 19 non-completers<br>Emotion dysregulation<br>78% female<br>9% Black, 6% Hispanic/Latine, 28% White, 4% multiracial, 15% other<br>$M$ age: 22.1 ($SD$ 4.8) - 22.3 ($SD$ 5.3) across groups | DBT-ST group vs. positive psychotherapy group<br>DBT-ST modules: all<br>DBT-ST sessions: 12 x 2 hours (some groups 11 sessions due to snow closures) | DBT-ST vs. positive psychotherapy: no significant between-group differences<br>Pre-post within groups: medium-to-large effect sizes for DBT-ST group (distress tolerance $d=.71$, emotion regulation $d=1.16$, mindfulness $d=1.07$) and small-to-medium effect sizes for positive psychotherapy group (distress tolerance $d=.38$, emotion regulation $d=.36$, mindfulness $d=.53$)<br>Acceptability: lower dropout rates for DBT-ST vs. positive psychotherapy. Average number of groups attended out of 12 for DBT-ST 9.04 ($SD$ 2.55) and for positive psychotherapy 5.31 ($SD$ 3.21). Working alliance inventory scores increased for both groups, with larger effect size for DBT-ST ($d=0.83$) vs. positive psychology ($d=0.69$) |
| Wieczorek et al. (2021)<br>Quasi-experimental<br>Australia (Sydney) | $N = 62$<br>ITT analysis with 21 non-completers<br>Emotion dysregulation<br>70% female completers ($n=33$), 91% female non-completers ($n=21$)<br>Race/ethnicity not reported<br>$M$ age: 33 ($SD$ 11.5) completers, 37 ($SD$ 14.2) non-completers | DBT-ST groups, pre-post, no comparison<br>DBT-ST modules: all<br>DBT-ST sessions: 15 x 2.5 hours (3 groups), 12 x 2.5 hours (4 groups), 11 x 2.5 hours (1 group, which omitted interpersonal effectiveness) | Pre vs. post: improvements in psychological distress ($d=0.851$, $p<.001$), days with disability/functional impairment ($d=0.638$, $p<.001$), depression ($d=0.605$, $p<.001$), and DBT skills use ($d=0.772$, $p<.001$) |
| Substance Use |  |  |  |
| Azizi et al. (2010)<br>Pilot RCT<br>Iran (Tehran) | $N = 39$<br>Competed case analysis<br>$N = 38$<br>Opioid use disorder<br>0% Female<br>Race/ethnicity not reported<br>$M$ age: 25.6 - 27.7 across groups | DBT-ST group vs. CBT group vs. medication only<br>DBT-ST modules: emotion regulation, mindfulness<br>DBT-ST and CBT sessions: 10 x 90 minutes each | DBT-ST and CBT vs. control: improvements in distress tolerance, emotion regulation, and drug use ($p<.05$), higher medication compliance ($\chi^2=17.71$, $p<.05$), and longer duration in treatment ($F=28.34$, $p<.01$)<br>DBT-ST vs. CBT: improvements in distress tolerance and emotion regulation ($p<.05$)<br>DBT-ST vs. CBT and control: improvements in relapse prevention (vs. CBT $p<.05$, vs. medication $p<.01$). Relapse rates for DBT-ST- 23%, CBT - 31%, control - 67%. |
| Basereh, Safarzadeh, & Hooman (2022)<br>RCT<br>Iran (Khuzestan) | *N* = 75<br># Non-completers not reported<br>Stimulant drug use disorder<br>Gender and race/ethnicity not reported<br>*M* age: 34.3 (*SD* 5.5) - 35.2 (*SD* 6.8) across groups | DBT-ST group vs. Structured Matrix Treatment (Matrix Institute, CA, US) vs. medication only<br>DBT-ST modules: distress tolerance, emotion regulation, mindfulness<br>DBT-ST sessions: 8 x 90 minutes each<br>Matrix sessions: 14 x 90 minutes each | DBT-ST *and* SMT vs. control: improvements in quitting self-efficacy, distress tolerance, and mindfulness ($p < .001$)<br>DBT-ST vs. SMT: improvements in distress tolerance ($p = .02$). No differences in quitting self-efficacy and mindfulness |
| Cooperman et al. (2019)<br>Quasi-experimental<br>United States (New Jersey) | *N* = 7<br>100% completion rate<br>Opioid use disorder, smoking<br>100% female<br>14% Hispanic/Latine, 86% White<br>*M* age: 39 (*SD* 14) | DBT group, pre-post, no comparison<br>DBT-ST modules: distress tolerance, emotion regulation, mindfulness<br>DBT modified for smoking cessation and drug relapse prevention as "DBT-Quit"<br>DBT sessions: 12 x 90 minutes each | Pre vs. post: smoked fewer cigarettes per day ($p < .05$), with lower CO levels ($p < .05$) and reduced nicotine dependence ($p < .05$)<br>Pre vs. post: no significant differences in emotion regulation, distress tolerance, or mindfulness skills |
| Edwards et al. (2022)<br>Quasi-experimental<br>United States (New York) | *N* = 13<br>Excludes 2 dropouts prior to study commencement<br>Substance use disorders, justice-involved<br>0% female<br>54% Black, 38% Hispanic/Latine, 8% White<br>*M* age: 44.4 (*SD* 11.5) | DBT-ST group, pre-post, no comparison<br>DBT-ST modules: all DBT-ST modified as DBT-J to include case management and criminal risk reduction for justice-involved veterans<br>DBT-ST sessions: 16 x 60-90 minutes each | Feasibility: Recruitment - 13 of 18 (72%) of potential participants approached successfully enrolled in program; Attendance/Retention - 10 of 13 (77%) completed program; Treatment Fidelity (adherence to treatment manual) $M = 4.8/5$ (*SD* 0.65)<br>Acceptability: Post-treatment, participants rated intervention $M = 22.14$ (*SD* 3.76) out of 25; Providers rated intervention as helpful ($M = 4.8/5$), beneficial to patients ($M = 4.8/5$), and relevant ($M = 4.6/5$)<br>Qualitative feedback from participants: appreciation of new skills, benefits of group setting, appreciation for providers, personal validation |
| Edwards et al. (2023)<br>Quasi-experimental<br>United States (New York) | *N* = 20<br>Non-completers = 3<br>Substance use disorders, justice-involved<br>5% female<br>55% Black, 30% Hispanic/Latine, 15% White<br>Age not reported | DBT-ST group, pre-post, no comparison<br>DBT-ST modules: all DBT-ST modified as DBT-J to include case management and criminal risk reduction for justice-involved veterans<br>DBT-ST sessions: 16 x 60-90 minutes each | Pre vs. post: improvement in psychological distress ($d = -0.46$), maintained at 1-month follow-up<br>Pre vs. post: reduction in alcohol use ($d = -0.88$) and non-alcohol substance use ($d = -0.41$), both maintained at 1-month follow-up |
| Nadimi & Pishgar (2015)<br>Pilot RCT<br>Iran (Khorasan) | *N* = 30<br># Non-completers not reported<br>Drug use disorders<br>100% female<br>Race/ethnicity not reported<br>*M* age: 29 (*SD* 6.8) | DBT-ST group vs. treatment-as-usual<br>DBT-ST modules: emotion regulation, interpersonal effectiveness, mindfulness<br>DBT-ST sessions: 20 x 90 minutes each | DBT-ST vs. treatment as usual: improvements in distress tolerance ($p = 0.000$), with results maintained at 2-month follow-up<br>Pre vs. post for DBT-ST group: improvements in all distress tolerance subscales (Tolerance, Attraction, Assessment, Regulation) ($p < .001$). No significant changes in subscales for treatment as usual group |
| Nyamathi et al. (2017)<br>RCT<br>United States (California) | *N* = 130<br>Completed case analysis<br>*N* = 116<br>Drug and alcohol use disorders, unhoused, justice-involved<br>100% female<br>37% - 45% Black, 40% Hispanic/Latine, 11% - 17% White, 5% - 6% other across groups<br>*M* age: 38.6 (*SD* 11.3) - 39.1 (*SD* 11.5) across groups | Individual and group sessions, DBT-ST vs. health promotion<br>DBT-ST modules: all<br>DBT-ST "corrections-modified" for parolees/probationers<br>DBT-ST and health promotion sessions: 16 x 2 hours | DBT-CM vs. health promotion: larger increase in 6-month drug use abstinence with urinalysis confirmation (*OR*=2.60, 95% CI [1.04, 6.53], *p*=.04) and on alcohol abstinence (*OR*=3.12, 95% CI [1.24, 7.85], *p*=.02). No difference for combined drug and alcohol abstinence<br>Multiple imputation analysis for DBT-CM vs. health promotion: no significant differences for 6-month abstinence<br>Multivariate logistic regression: lower odds of drug use abstinence for Black, Hispanic/Latine, and other race/ethnicity vs. White: Black (*aOR*=0.05, 95% CI [0.01.0.50], *p*=.01), Hispanic/Latine (*aOR*=0.08, 95% CI [0.01.0.74], *p*=.03), other (*aOR*=0.05, 95% CI [0.00.0.64], *p*=.02) |
| Rezaie, Afshari, & Balagabri (2021)<br>Pilot RCT<br>Iran (Isfahan) | *N* = 50<br>ITT analysis with 2 non-completers<br>Opioid use disorder<br>0% female<br>Race/ethnicity not reported<br>*M* age: 34.1 (*SD* 5) - 36 (*SD* 4.1) across groups | DBT-ST + methadone group vs. medication only<br>DBT-ST modules: distress tolerance, emotion regulation, mindfulness<br>DBT-ST sessions: 16 x 90 minutes | DBT-ST + methadone vs. methadone only: at 3-month follow-up, improvements in distress tolerance (*p*<0.001), emotion regulation (*p*<0.001), craving (*p*<0.001), and depression (*p*=0.002) |
| Sahranavard & Miri (2018)<br>Quasi-experimental<br>Iran (South Khorasan) | *N*=30<br>Substance use disorders<br>100% female<br>Race/ethnicity not reported<br>*M* age=34.1, age range: 25-40 | DBT-ST group vs. CBT group vs. waitlist control<br>DBT-ST modules: all<br>DBT-ST and CBT sessions: 8 x 90 minutes each | DBT-ST and CBT vs. control: improvements in depression symptoms for DBT-ST (*p*<0.001, *η*<sup>2</sup>=0.86) and CBT (*p*<0.001, *η*<sup>2</sup>=0.75) |

### Study Characteristics

Of the 23 unique research studies in this review, 12 were quasi-experimental designs, seven were pilot RCTs, and four were full RCTs. Given the low number of full RCTs, we did not perform a systematic quality assessment of studies. Eleven studies were conducted in Iran, nine in the United States, two in Australia, and one in Canada. The target populations were patients with breast cancer,^27,41^ people with emotion dysregulation,^30,38–40,42–52^ and people with substance use problems.^29,31,32,34,53–57^ The three Australian and Canadian studies examined only emotion dysregulation. The Iranian and U.S. studies included all three populations types– breast cancer, emotion dysregulation and substance use.

Study sample sizes ranged from 7 to 130 participants (*M* 42.22, *SD* 27.52). Participant gender was classified as male/female, with one study not reporting gender ^34^. Over half (59%) of the 22 studies reporting gender focused on a single gender. Nine studies included female-only populations and four studies were male-only. All but one^42^ of the 11 Iranian studies were single-gender, while the three studies in Australia and Canada were mixed-gender. The nine U.S. studies included each type, with three single-gender and the rest mixed-gender. Both breast cancer study samples were female. Three emotion dysregulation studies focused on females and one study on males. The remaining eight studies of emotion dysregulation had mixed-gender populations that were predominantly female at 59% - 94% of participants. With one exception among the substance use studies, a study with 5% female participation ^55^, study samples were only female (four studies) or only male (three studies).

Ten studies reported race/ethnicity composition – eight studies from the U.S. and one each from Canada and Iran. Most (80%) of these studies reported non-White participation rates approaching, at, or above national norms, ranging from 14% to 92% of study samples. Sixteen studies reported mean ages, showing that all but one study^46^ were conducted in relatively young populations ranging from mean ages of 21.3 – 44.4 years. Of the 11 studies reporting age ranges, only four included any participants over age 50.^27,30,46,49^

### Intervention Structure

All DBT-ST brief interventions were administered as group sessions, with two studies alternating group and individual DBT-ST sessions.^27,31^ The number of sessions ranged from 4 to 20, with an average of 11.65 sessions (*SD* 4.07). Over half of studies (57%) reported on professions of group facilitators, which included doctoral and master’s level mental health clinicians and trainees, nurses, and community health workers. Nine of these studies noted that group facilitators were specifically trained in DBT and/or DBT-ST, with training periods ranging from 10 hours to 10 days. Facilitator experience leading DBT-ST groups ranged from none to 15 years.

### Intervention Content

Two-thirds of studies included all four modules of DBT-ST: mindfulness, emotion regulation, distress tolerance, and interpersonal effectiveness. Five studies excluded interpersonal effectiveness^32,34,47,50,56^ and one study excluded distress tolerance,^29^ while retaining the other three modules. The remaining two studies included emotion regulation and either distress tolerance^53^ or mindfulness.^48^ The number of modules covered and intervention length were unrelated, as all four modules appeared in interventions as short as four weeks.^47^

For intervention development, most studies (87%) drew from DBT training manuals, books, and/or seminal articles by the DBT founder, Marsha Linehan.^25,58^ Three studies did not cite Linehan DBT literature.^27,46,57^ Of studies that published individual session outlines with their articles, nine appeared to use published DBT-ST materials with minimal modification, including one study that did not cite the Linehan source material.^57^ Three studies adapted DBT-ST to include concepts and materials specific to their populations and created unique names for these interventions: DBT-Quit for cigarette smoking,^32^ DBT-J for “Justice”,^54,55^ and DBT-CM for “Case Management”.^31^

### Comparison Populations

Eleven studies were pilot or full RCTs with one or two comparison populations per study. Seven of these RCTs included medication-only,^34,53,56^ treatment-as-usual,^29,41^ or waitlist^43,45^ control groups. Other RCT comparators were cognitive behavioral therapy^42,53^ and structured programs or educational groups.^30,31,34,51^

### Measures

Measurement instruments appearing in studies in this review are shown in Table 2 Measures. Emotion regulation and mindfulness were measured by at least two different instruments. Distress tolerance was measured by one instrument. Eight different instruments measured states of psychological distress. Another commonly appearing measure captured frequencies of DBT skills use. There were no instruments to measure interpersonal effectiveness, as assessments for this construct have yet to be developed.

**Table 2.** Measures.

| DBT Target | Measure | Description | Study Inclusion |
| --- | --- | --- | --- |
| DBT Skills Use | DBT Ways of Coping Checklist (DBT-WCCL) | 59-item self-report of DBT skills use | 30,48,51,52 |
| Distress Tolerance | Distress Tolerance Scale (DTS) | 15-item self-report of negative beliefs about feeling distressed or upset | 29,32,34,51,53,56 |
| Emotion Regulation | Difficulty in Emotion Regulation Scale (DERS) | 36-item self-report of six facets of emotion dysregulation | 30–32,42–44,46–48,51,53 |
| Emotion Regulation | Emotional Regulation Questionnaire | 10-item self-report of habitual use of emotional regulation strategies | 56 |
| Emotion Regulation | Positive and Negative Affect Schedule (PANAS-X) | 60-item self-report of higher-order positive and negative emotions | 48 |
| Emotion Regulation | Emotion Stability Scale from International Personality Item Pool | 20-item self-report of positive and negative emotions | 49 |
| Mindfulness | Kentucky Inventory of Mindfulness Skills (KIMS) | 39-item self-report of mindfulness practices (aka Five Facet Mindfulness Questionnaire) | 32,40,42,47,48,51 |
| Mindfulness | Mindful Attention Awareness Scale (MAAS) | 15-item self-report of mindful awareness | 34,49 |
| Psychological Distress | Brief Symptom Inventory-18 (BSI-18) | 18-item self-report of psychological distress – somatization, depression, and anxiety | 27 |
| Psychological Distress | Counseling Center Assessment of Psychological Symptoms-34 (CCAPS-34) | 34-item self-report of psychological symptoms in students | 47 |
| Psychological Distress | Depression Anxiety Stress Scales (DASS) | 42-item self-report of depression, anxiety, and stress/tension | 27,48 |
| Psychological Distress | Distress Scale | Visual analog scale self-rating for any type of psychological distress | 27 |
| Psychological Distress | General Health Questionnaire-12 (GHQ-12) | 12-item self-report of psychological distress indicative of mental health concerns | 49,55 |
| Psychological Distress | General Health Questionnaire-28 (GHQ-28) | 28-item self-report of psychological distress indicative of mental health concerns | 53 |
| Psychological Distress | Kessler Psychological Distress Scale (K-10) | 10-item self-report of psychological distress over the past month | 41,52 |
| Psychological Distress | Outcome Questionnaire-45 (OC-45) | 45-item self-report of problems leading to distress in symptoms, relationships, roles | 30 |

The lack of alignment between the outcome measures and the DBT-ST constructs included in individual studies is notable. All studies included emotion regulation and mindfulness in DBT-ST sessions, but only half measured emotion regulation and just one-fourth measured mindfulness. Nearly all studies included distress tolerance in sessions, with only six measuring it.

### Primary Outcomes

Results from every study showed a significant positive change in at least one primary outcome associated with a DBT-ST intervention. The most frequently measured constructs and most common improvements were in distress tolerance skills and/or psychological distress, reported by 14 studies. Eleven studies measured and found significant improvements in emotion regulation. Participants in six of eight studies measuring mindfulness improved these skills. The four studies that tracked substance use, either by self-report or biochemical verification, found that DBT-ST led to reduced use or abstinence, including for cigarette smoking. Studies examining the feasibility and/or acceptability of DBT-ST found both, with good recruitment and retention rates and participant satisfaction with the intervention. Of the eight studies incorporating one- to six-month follow-up periods, all but one found maintenance or improvement at follow-up in at least one post-treatment outcome.

However, when DBT-ST was compared to another evidence-based treatment, such as cognitive behavioral therapy or an established substance use treatment program, there were few, if any, between-group differences; significant differences were limited to changes in DBT constructs, such as emotion regulation and distress tolerance. Overall, DBT-ST was not found to be inferior to other treatments, but also not clearly superior.

### Outcomes by Condition

#### Cancer

The DBT-ST interventions in both studies of breast cancer were among the shortest in this review, at 8 sessions^27^ and 10 sessions.^41^ Both interventions resulted in significant decreases in distress levels, with Faraj^41^ finding that the intervention explained nearly 50% of the decrease. Participants in Anderson et al.^27^ reported improved ability to cope, recognize stress triggers, and use DBT-ST skills, while participants in Faraj^41^ became more hopeful that they would live longer.

#### Emotion Dysregulation

In addition to improvements in DBT-related targets such as emotion regulation and distress tolerance, DBT-ST interventions led to reductions in depression and anxiety in each of the seven studies that measured one or both conditions. Reductions were maintained for the six studies that followed participants for one to six months. In the only study of older adults, with mean age 63, Khodabakhshi-Kooleaa et al.^46^ found that DBT-ST participants gained abilities to manage emotions and reduced their anxiety about death. In a study of college students, Lee & Arora^47^ succeeded in developing and testing a short 4-week DBT-ST program. The students were laudatory of the program, with 92% attending at least three of the four sessions and 91% reporting improved ability to cope with adversity, while improving skills in emotion regulation and mindfulness, increasing resilience, and reducing distress. In Uliaszek et al.,^51^ DBT-ST was compared to positive psychotherapy, with the DBT-ST participants noting stronger therapeutic working alliances with facilitators and lower likelihood of dropping out of treatment. Wieczorek et al.,^52^ who conducted a pre-post examination of eight “real-world” DBT-ST groups, reported decreases in depression and a decrease in days with functional impairment.

#### Substance Use

For outcomes specific to substance use, participants reduced drug use, relapses, and medication non-compliance,^53^ increased quitting self-efficacy,^34^ and reduced cravings and depression.^56^ A small study of women receiving methadone treatment who also smoked cigarettes found that DBT-ST helped them reduce use from a median of 12 to 2 cigarettes per day.^32^ Three studies of DBT-ST for substance use were with justice-involved individuals not in treatment.^31,54,55^ Nyamathi et al.^31^ conducted a full RCT with an unhoused, formerly incarcerated female population engaging in alcohol and drug use. While completed case analysis (*N*=116) showed large reductions in substance use at six months, these differences did not hold for multiple imputation analysis with the entire sample (*N*=130). In Edwards et al.,^54^ U.S. veterans rated the DBT-ST intervention highly, appreciated acquiring new skills, and found comfort and validation in the group setting. In a follow-on study, veterans were able to reduce alcohol and drug use as well as psychological distress, changes that were maintained at one-month follow-up.^55^

## Discussion

We conducted this review to evaluate the suitability of DBT-ST as a brief intervention for cigarette smoking by patients with cancer. We located and synthesized 26 publications describing 23 unique studies of DBT-ST in populations similar to our target population, specifically people with substance use problems, people with difficulties in emotion regulation, and patients with cancer. DBT-ST as a standalone treatment in an abbreviated format is relatively recent, so many studies were exploratory, with considerable heterogeneity in intervention design, testing, and measurement. Of the studies reporting race/ethnicity, most recruited diverse participants, with some studies far exceeding national population averages. All studies found at least one improvement in a condition or behavior following DBT-ST intervention, with improvements maintained at follow-up. However, DBT-ST’s superiority when compared to other active group treatments was not established.

Our results are similar to those found by Valentine et al.^59^ in their review of DBT-ST as a standalone treatment, which included interventions of greater length and for a wider set of conditions than this review. Their review of 31 studies found consistent effectiveness for DBT-ST across conditions ranging from depression and suicidality to eating disorders, but no significant differences between DBT-ST and other manualized treatments. Studies varied greatly in design, target population, intervention adaptations, selected measurements, and outcomes of interest. Our results are also similar to a systematic literature review of DBT-ST for substance use disorders by Warner & Murphy ^33^, which included studies with more intensive and longer interventions, with some taking place in inpatient settings. The nine studies in this review differed greatly from each other in research design and quality and in how DBT-ST was adapted for target populations. Yet together they provided preliminary support for DBT-ST as an effective standalone intervention for substance use disorders.

## Implications for Clinical Practice and Research

DBT-ST, which has been employed in clinical practice for many decades as part of DBT programs, is now emerging as a promising brief intervention for conditions characterized by difficulties managing stress and regulating emotions, such as cancer and smoking. A strength across studies was the inclusion of diverse racial and ethnic populations, which Harned, Coyle, & Garcia^28^ also found. However, DBT-ST research is in its nascent period and improvement in the rigor of future studies is warranted. First, the paucity of RCTs and/or lack of comparison groups, particularly active treatment groups, prevented causal inference of brief DBT-ST as an independent predictor of change. Future studies should prioritize intervention development pathways that lead to RCTs, such as outlined by Rounsaville, Carroll, & Onken.^60^ Second, with one exception,^54,55^ each study was the sole DBT-ST study by the research group, meaning further testing of interventions either was not performed or not published. Third, females and younger adults were over-represented in the studies; researchers should conduct intentional recruitment and enrollment of male and older adult participants in DBT-ST research. Fourth, greater transparency about intervention design would enhance replicability. Fifth, studies exhibited little agreement regarding what outcomes should be measured and how. Often the major targets of interventions were not measured, notably emotion regulation, distress tolerance, mindfulness, interpersonal effectiveness and condition-specific outcomes such as substance use. Measurement instruments selected by studies varied widely, reducing comparability of results across studies. Finally, only about one-fourth of studies followed up with participants after intervention completion, so that little is known about the longer-term effects of brief DBT-ST interventions. In summary, the implications for future DBT-ST brief intervention research include prioritization of RCTs, continuity of intervention research, recruitment of male and older adult participants, transparency about intervention design, appropriate selection of outcomes, and follow-up.

## Limitations

Our results may be limited by several factors. Most studies in this review were quasi-experimental or pilot RCT studies, so the preliminary findings may not hold for future studies. We allowed all quantitative study designs due to the early nature of DBT-ST research, so we often lacked comparison groups and adequate sample sizes to increase confidence in our findings. We did not conduct a formal quality review of studies due to the lack of RCTs, so did not assess and compare critical features of studies that may have raised further doubts or increased confidence in results.

## Conclusion

We conclude from this review that DBT-ST holds promise as an adapted intervention for cigarette smoking by patients diagnosed with cancer. Preliminary results indicate that DBT-ST may be effective for similar populations, such as patients with breast cancer, people having difficulty regulating emotions, and people with substance use problems. DBT-ST can be delivered in as few as four sessions, which is more suitable for patients with cancer who may have time-intensive treatment schedules. DBT-ST appears to be feasible and acceptable for diverse populations. However, DBT-ST study samples are mostly female and younger, whereas patients with cancer who continue to smoke tend to be male and older. Despite this concern, the preliminary results are convincing enough to pursue adapting DBT-ST as a brief intervention for patients with cancer who smoke cigarettes.

## Data Availability

All data produced in the present work are contained in the manuscript

## Author contributions using CRediT

Marcia H. McCall: conceptualization, data curation, formal analysis, investigation, methodology, project administration, resources, visualization, writing – original draft, writing – review and editing

Charlotte T. Boyd: data curation, investigation, project administration, resources, visualization, writing – original draft, writing – review and editing

Nicole D. Kerr: writing – original draft, writing – review and editing

Erin L. Sutfin: supervision, writing – review and editing

Stephanie S. Daniel: supervision, writing – review and editing

## Notes

### Competing Interest Statement

The authors have declared no competing interest.

### Funding Statement

This study did not receive any funding

## REFERENCES

1. US Office on Smoking and Health. The Health Consequences of Smoking—50 Years of Progress: A Report of the Surgeon General. Centers for Disease Control and Prevention (US); 2014.

2. US Office on Smoking and Health. Smoking Cessation: A Report of the Surgeon General. Centers for Disease Control and Prevention (US); 2020:700. https://stacks.cdc.gov/view/cdc/84557

3. Lucchiari C, Masiero M, Botturi A, Pravettoni G. Helping patients to reduce tobacco consumption in oncology: a narrative review. SpringerPlus. 2016;5(1):1–18. doi:10.1186/s40064-016-2798-9

4. Caini S, Del Riccio M, Vettori V, et al. Quitting smoking at or around diagnosis improves the overall survival of lung cancer patients: a systematic review and meta-analysis. J Thorac Oncol. 2022;17(5):623–636. doi:10.1016/j.jtho.2021.12.005

5. Puleo GE, Borger T, Bowling WR, Burris JL. The state of the science on cancer diagnosis as a “teachable moment” for smoking cessation: a scoping review. Nicotine Tob Res. 2022;24(2):160–168. doi:10.1093/ntr/ntab139

6. Westmaas JL, Newton CC, Stevens VL, Flanders WD, Gapstur SM, Jacobs EJ. Does a recent cancer diagnosis predict smoking cessation? An analysis from a large prospective US cohort. J Clin Oncol. 2015;33(15):1647–1652. doi:10.1200/JCO.2014.58.3088

7. National Cancer Center Network. NCCN Smoking Cessation Guidelines Detail. NCCN. August 30, 2022. Accessed November 6, 2022. https://www.nccn.org/guidelines/guidelines-detail

8. Sheeran P, Jones K, Avishai A, et al. What works in smoking cessation interventions for cancer survivors? A meta-analysis. Health Psychol. 2019;38(10):855–865. doi:10.1037/hea0000757

9. Vinci C. Cognitive behavioral and mindfulness-based interventions for smoking cessation: a review of the recent literature. Curr Oncol Rep. 2020;22(6). doi:10.1007/s11912-020-00915-w

10. Duffy SA, Louzon SA, Gritz ER. Why do cancer patients smoke and what can providers do about it? Community Oncol. 2012;9(11):344–352. doi:10.1016/j.cmonc.2012.10.003

11. Wells M, Aitchison P, Harris F, et al. Barriers and facilitators to smoking cessation in a cancer context: a qualitative study of patient, family and professional views. BMC Cancer. 2017;17. doi:10.1186/s12885-017-3344-z

12. Berg CJ, Haardörfer R, McBride CM, et al. Resilience and biomarkers of health risk in Black smokers and nonsmokers. Health Psychol Off J Div Health Psychol Am Psychol Assoc. 2017;36(11):1047–1058. doi:10.1037/hea0000540

13. Hughes K, Bellis MA, Hardcastle KA, et al. The effect of multiple adverse childhood experiences on health: a systematic review and meta-analysis. Lancet Public Health. 2017;2(8):e356–e366. doi:10.1016/S2468-2667(17)30118-4

14. Llabre MM, Schneiderman N, Gallo LC, et al. Childhood Trauma and Adult Risk Factors and Disease in Hispanics/Latinos in the US: Results From the Hispanic Community Health Study/Study of Latinos (HCHS/SOL) Sociocultural Ancillary Study. Psychosom Med. 2017;79(2):172–180. doi:10.1097/PSY.0000000000000394

15. Merrick MT, Ford DC, Ports KA, et al. Vital Signs: Estimated Proportion of Adult Health Problems Attributable to Adverse Childhood Experiences and Implications for Prevention - 25 States, 2015-2017. MMWR Morb Mortal Wkly Rep. 2019;68(44):999–1005. doi:10.15585/mmwr.mm6844e1

16. Ports KA, Holman DM, Guinn AS, et al. Adverse Childhood Experiences and the Presence of Cancer Risk Factors in Adulthood: A Scoping Review of the Literature From 2005 to 2015. J Pediatr Nurs. 2019;44:81–96. doi:10.1016/j.pedn.2018.10.009

17. Cludius B, Mennin D, Ehring T. Emotion regulation as a transdiagnostic process. Emotion. 2020;20(1):37–42. doi:10.1037/emo0000646

18. Guimond AJ, Ivers H, Savard J. Is emotion regulation associated with cancer-related psychological symptoms? Psychol Health. 2019;34(1):44–63. doi:10.1080/08870446.2018.1514462

19. Smith IS, Wellecke C, Weihs KL, Bei B, Wiley JF. Piloting CanCope: An internet-delivered transdiagnostic intervention to improve mental health in cancer survivors. Psychooncology. 2022;31(1):107–115. doi:10.1002/pon.5787

20. Vaughan E, Koczwara B, Kemp E, Freytag C, Tan W, Beatty L. Exploring emotion regulation as a mediator of the relationship between resilience and distress in cancer. Psychooncology. 2019;28(7):1506–1512. doi:10.1002/pon.5107

21. Lee J, Cheong J, Markham MJ, Lam J, Warren GW, Salloum RG. Negative affect and the utilization of tobacco treatment among adult smokers with cancer. Psychooncology. 2021;30(1):93–102. doi:10.1002/pon.5543

22. Schlam TR, Baker TB, Smith SS, Cook JW, Piper ME. Anxiety Sensitivity and Distress Tolerance in Smokers: Relations With Tobacco Dependence, Withdrawal, and Quitting Success†. Nicotine Tob Res. 2020;22(1):58–65. doi:10.1093/ntr/ntz070

23. U.S. National Cancer Institute. Treating Smoking in Cancer Patients: An Essential Component of Cancer Care. National Cancer Institute Tobacco Control Monograph 23. U.S. Department of Health and Human Services, National Institutes of Health, National Cancer Institute; 2022:325.

24. Dimeff LA, Linehan MM. Dialectical behavior therapy for substance abusers. Addict Sci Clin Pract. 2008;4(2):39–47.

25. Linehan MM. Dialectical Behavioral Therapy Skills Training Manual. The Guilford Press; 2015.

26. Pietrek C, Elbert T, Weierstall R, Müller O, Rockstroh B. Childhood adversities in relation to psychiatric disorders. Psychiatry Res. 2013;206(1):103–110. doi:10.1016/j.psychres.2012.11.003

27. Anderson RC, Jensik K, Peloza D, Walker A. Use of the dialectical behavior therapy skills and management of psychosocial stress with newly diagnosed breast cancer patients. Plast Surg Nurs Off J Am Soc Plast Reconstr Surg Nurses. 2013;33(4):159–163. doi:10.1097/PSN.0000000000000018

28. Harned MS, Coyle TN, Garcia NM. The inclusion of ethnoracial, sexual, and gender minority groups in randomized controlled trials of dialectical behavior therapy: a systematic review of the literature. Clin Psychol Sci Pract. 2022;29(2):83–93. doi:10.1037/cps0000059

29. Nadimi M, Pishgar M. Effectiveness of dialectical behavioral therapy in increasing distress tolerance of women drug abusers. 2015;5:844–849.

30. Neacsiu AD, Eberle JW, Kramer R, Wiesmann T, Linehan MM. Dialectical behavior therapy skills for transdiagnostic emotion dysregulation: a pilot randomized controlled trial. Behav Res Ther. 2014;59:40–51. doi:10.1016/j.brat.2014.05.005

31. Nyamathi AM, Shin SS, Smeltzer J, et al. Achieving drug and alcohol abstinence among recently incarcerated homeless women: a randomized controlled trial comparing dialectical behavioral therapy-case management with a health promotion program. Nurs Res. 2017;66(6):432–441. doi:10.1097/NNR.0000000000000249

32. Cooperman NA, Rizvi SL, Hughes CD, Williams JM. Field test of a dialectical behavior therapy skills training-based intervention for smoking cessation and opioid relapse prevention in methadone treatment. J Dual Diagn. 2019;15(1):67–73. doi:10.1080/15504263.2018.1548719

33. Warner N, Murphy M. Dialectical behaviour therapy skills training for individuals with substance use disorder: a systematic review. Drug Alcohol Rev. 2022;41(2):501–516. doi:10.1111/dar.13362

34. Basereh S, Safarzadeh S, Hooman F. The effectiveness of group dialectical behavior therapy and structured matrix treatment on quit addiction self-efficacy, distress tolerance, and mindfulness in individuals with stimulant drug abuse. J Health Rep Technol. 2022;8(4):e127427. doi:10.5812/jhrt-127427

35. Mak S, Thomas A. Steps for Conducting a Scoping Review. J Grad Med Educ. 2022;14(5):565–567. doi:10.4300/JGME-D-22-00621.1

36. Tricco AC, Lillie E, Zarin W, et al. PRISMA Extension for Scoping Reviews (PRISMA-ScR): Checklist and Explanation. Ann Intern Med. 2018;169(7):467–473. doi:10.7326/M18-0850

37. Covidence systematic review software. Published online 2024. www.covidence.org

38. Neacsiu AD, Rompogren J, Eberle JW, McMahon K. Changes in Problematic Anger, Shame, and Disgust in Anxious and Depressed Adults Undergoing Treatment for Emotion Dysregulation. Behav Ther. 2018;49(3):344–359. doi:10.1016/j.beth.2017.10.004

39. Southward MW, Eberle JW, Neacsiu AD. Multilevel associations of daily skill use and effectiveness with anxiety, depression, and stress in a transdiagnostic sample undergoing dialectical behavior therapy skills training. Cogn Behav Ther. 2022;51(2):114–129. doi:10.1080/16506073.2021.1907614

40. Wyatt KP, Eberle JW, Ruork AK, Neacsiu AD. Mechanisms of change in treatments for transdiagnostic emotion dysregulation: The roles of skills use, perceived control and mindfulness. Clin Psychol Psychother. 2023;30(6):1380–1392. doi:10.1002/cpp.2879

41. Faraji M. The effectiveness of dialectical behavior therapy (DBT) in reducing distress and increased life expectancy in patients with breast cancer. Cumhur Üniversitesi Fen Edeb Fakültesi Fen Bilim Derg. 2015;36(3):1340–1346.

42. Afshari B, Hasani J. Study of Dialectical Behavior Therapy Versus Cognitive Behavior Therapy on Emotion Regulation and Mindfulness in Patients with Generalized Anxiety Disorder. J Contemp Psychother. 2020;50(4):305–312. doi:10.1007/s10879-020-09461-9

43. Babaheydari SHM, Homayooni R, Zare R, Giski MM, Khodarahimi S, Rasti A. The effectiveness of group-based dialectical behavior therapy on emotional regulation problems and anxiety strictness in males with generalized anxiety disorder. Curr Psychol J Diverse Perspect Diverse Psychol Issues. 2024;43(20):18253–18261. doi:10.1007/s12144-024-05666-6

44. Davarani ZZ, Heydarinasab L. The effectiveness of dialectical behavior therapy skills to reduction in difficulty in emotion regulation among students. Rev Argent Clínica Psicológica. 2019;28(5):842–848.

45. Harley R, Sprich S, Safren S, Jacobo M, Fava M. Adaptation of dialectical behavior therapy skills training group for treatment-resistant depression. J Nerv Ment Dis. 2008;196(2):136–143. doi:10.1097/NMD.0b013e318162aa3f

46. Khodabakhshi-Koolaee A, Falsafinejad MR, Zoljalali T, Ghazizadeh C. Dialectical Behavior Therapy: Effect on Emotion Regulation and Death Anxiety in Older Female Adults. Omega J Death Dying. 2024;88(3):1218–1231. doi:10.1177/00302228211065960

47. Lee S, Arora IS. The effectiveness, acceptability, and sustainability of a 4-week DBT-informed group therapy in increasing psychological resilience for college students with mental health issues. J Clin Psychol. 2023;79(9):1929–1942. doi:10.1002/jclp.23509

48. Rizvi SL, Steffel LM. A Pilot Study of 2 Brief Forms of Dialectical Behavior Therapy Skills Training for Emotion Dysregulation in College Students. J Am Coll Health. 2014;62(6):434–439. doi:10.1080/07448481.2014.907298

49. Robins TG, Roberts RM, Sarris A. The effectiveness, feasibility, and acceptability of a dialectical behaviour therapy skills training group in reducing burnout and psychological distress in psychology trainees: A pilot study. Aust Psychol. 2019;54(4):292–301. doi:10.1111/ap.12389

50. Shojaei-Fadafen H, Jajarmi M, Mahdian H. Evaluation of the Effectiveness of Dialectical Behavior Therapy on Depressive Symptoms, Irrational Beliefs and Psychological Well-being of Women. Int J Behav Sci. 2022;15(4):287–293. doi:10.30491/IJBS.2022.279258.1515

51. Uliaszek AA, Rashid T, Williams GE, Gulamani T. Group therapy for university students: A randomized control trial of dialectical behavior therapy and positive psychotherapy. Behav Res Ther. 2016;77:78–85. doi:10.1016/j.brat.2015.12.003

52. Wieczorek M, Kacen T, King B, Wilhelm K. The effectiveness of a short-term DBT skills group in a ‘real-world’ clinical setting. Australas Psychiatry. 2021;29(6):600–603. doi:10.1177/10398562211038907

53. Azizi A, Borjali A, Golzari M. The Effectiveness of Emotion Regulation Training and Cognitive Therapy on the Emotional and Addictional Problems of Substance Abusers. Iran J Psychiatry. 2010;5(2):60–65.

54. Edwards ER, Dichiara A, Epshteyn G, et al. Dialectical behavior therapy for justice-involved veterans (DBT-J): Feasibility and acceptability. Psychol Serv. 2022;20(Suppl 2):108–121. doi:10.1037/ser0000691

55. Edwards ER, Epshteyn G, Snyder S, et al. Dialectical behavior therapy for justice-involved veterans: Changes in treatment targets in a small, pre-post design clinical trial. Psychol Serv. 2023;20(Suppl 2):98–107. doi:10.1037/ser0000766

56. Rezaie Z, Afshari B, Balagabri Z. Effects of Dialectical Behavior Therapy on Emotion Regulation, Distress Tolerance, Craving, and Depression in Patients with Opioid Dependence Disorder. J Contemp Psychother. Published online February 4, 2021. doi:10.1007/s10879-020-09487-z

57. Sahranavard S, Miri MR. A comparative study of the effectiveness of group-based cognitive behavioral therapy and dialectical behavioral therapy in reducing depressive symptoms in Iranian women substance abusers. Psicol Reflex E Crítica. 2018;31(1):15. doi:10.1186/s41155-018-0094-z

58. Linehan MM. Cognitive-Behavioral Treatment of Borderline Personality Disorder. Guilford Publications; 1993. http://ebookcentral.proquest.com/lib/wfu/detail.action?docID=330598

59. Valentine SE, Smith AM, Stewart K. Chapter 15 - A review of the empirical evidence for DBT skills training as a stand-alone intervention. In: Bedics J, ed. The Handbook of Dialectical Behavior Therapy. Academic Press; 2020:325–358. doi:10.1016/B978-0-12-816384-9.00015-4

60. Rounsaville BJ, Carroll KM, Onken LS. A stage model of behavioral therapies research: getting started and moving on from stage I. Clin Psychol Sci Pract. 2001;8(2):133–142. doi:10.1093/clipsy.8.2.133

